# Age-appropriate Vaccination and Associated Factors among Children Aged 12- 35 Months in Ethiopia: A Multi-Level Analysis

**DOI:** 10.1101/2024.06.06.24308578

**Authors:** Bekelu Teka Worku, Eshetu Alemayehu Wordofa, Gadisa Senbeto, Beakal Zinab, Ebissa Bayana Kebede, Fira Abamecha, Gurmessa Tura Debela, Negalign Birhanu, Yibeltal Siraneh, Dessalegn Tamiru

**Author notes:** **Corresponding Author** (EBK).

## Abstract

**Background:** Age-appropriate vaccination is one of the key public health measures to prevent morbidity and mortality worldwide. Despite its importance, there has been insufficient emphasis on tackling this problem. Therefore, this study aimed to determine the prevalence of age-appropriate vaccination and associated factors in Ethiopia.

**Method:** Data of 1077 children aged 12-35months were extracted from the Ethiopian Mini Demographic and Health Survey 2019 using a prepared data extraction checklist and included in the analysis. The extracted data was analyzed using STATA version 14.0. Descriptive and inferential statistics were applied. then analysis at different levels was done using multilevel logistic regression. Significant variables were identified at p-value < 0.05 within 95% confidence level and AOR.

**Result:** The pooled prevalence of age-appropriate vaccination in this study was 21.17% at 95%CI (18.73-23.61). Factors like mothers aged >= 40 (AOR=4.05 at 95%CI1.03, 15.83), 35-39 years (AOR= 4.62 at 95%CI1.27,16.71), 25-29 years, (AOR =4.07 at 95%CI 1.18,14.03), Maternal secondary education (AOR=1.85 at 95% CI 1.06, 3.22), Maternal primary education (AOR= 1.60 at 95% CI1.07, 2.41) and rural residence (AOR=0.34, 95%CI 0.23,0.51) were significant predictors of age-appropriate vaccinations.

**Conclusion:** This study concluded that the prevalence of age-appropriate vaccination of children in Ethiopia is below the desired level. Hence, the stakeholders should give due attention to the timely vaccination of children equally as the effort being made to increase the coverage.

## Introduction

Vaccination involves exposing children to a weakened or killed version of a pathogen. This provokes a response in the immune system without causing serious infections (1). Immunization is a crucial public health intervention that can help to reduce child morbidity and mortality rates and increase life expectancy (2). The most common vaccine-preventable diseases among children include measles, diphtheria, influenza, tetanus, pertussis, hepatitis, mumps, pneumonia, polio, rotavirus, and others (3, 4).

To fight those vaccine-preventable diseases, the Expanded Program on Immunization (EPI) was launched by the World Health Organization (WHO) in 1974 with the main aim of providing immunization to all children worldwide. In 1984, the WHO standardized a vaccination schedule for six vaccines, namely Bacillus Calmette-Guérin (BCG), diphtheria-tetanus-pertussis (DTP), oral polio, and measles (5, 6). Similarly, Ethiopia launched an immunization program in 1980 as the EPI. The EPI was established to achievea 100% immunization rate in 10 years even though the original target remained unachieved after 20 years also (7, 8).

It is crucial to ensure that children receive vaccinations for their age to safeguard them from vaccine-prevalent diseases (9). Moreover, ensuring universal vaccination through routine and catch-up vaccination schedules is an essential part of quality healthcare in nations. It is linked to improved health outcomes, cognitive development, productivity, and cost savings (10–12). In line with this, the World Immunization Agenda 2030 recommends leaving no one behind for vaccination through increasing equitable access and use of new and existing vaccines (13). However, the burden of vaccine-preventable disease remains not easy in today’s world.

Nowadays, full vaccination of children timely at their appropriate age is a global public problem, and vaccination coverage has fallen back or stagnated (14). Timely vaccination among children ranges from 38% to 65% in the Philippines and median on-time vaccination in Sub-Saharan African countries is less than 50% (15, 16). In Ethiopia, the age-appropriate vaccination practice is less than half 43.7%, in the Afar region, nearly one-third 33.7% in Oromia Region, and 39.1% for specific vaccines in the North East Ethiopia (17–19). However, the pooled full vaccination coverage in Ethiopia is greater than 65% which is higher than all these figures indicating poor age-appropriate vaccination in different regions (20). On the other way, the magnitude of missed opportunities for routine vaccination is high 39.8% in Ethiopia despite the high value ofage-appropriatevaccinations(21).

Despite remarkable improvements in child health services utilization, age-appropriate childhood immunization is still challenging in Ethiopia (22). To avert this Ethiopia developed strategies and policies likethe Health Sector Transformation Plan (HSTP), reaching every district (RED) and sustainable outreach service (SOS) approaches and comprehensive multiyear immunization plans (cMYP)(23, 24). In general, the majority of the vaccination types are age-specific and some vaccinations could not be recommended after some point of time in children’s age. It is highly important to comply with age-appropriate vaccinations to provide maximum effectiveness against vaccine-preventable diseases among children (9).

Revealing information on the age appropriateness of vaccination from national data can help to provide a more complete view of vaccination coverage in the country. The information could be essential to clarify the missing and delayed vaccination status that the stakeholders such as policymakers can utilize to address the gaps in immunization programs. Although some studies have assessed age-appropriate vaccination in Ethiopia, no study used the most recent national data to evaluate this issue. Therefore, this study aimed to conduct a multi-level analysis to determine the level of age-appropriate vaccination status and associated factors in Ethiopia using the 2019 Ethiopia Mini Demographic and Health Survey (25).

## Materials and Methods

### Source of data and study design

The study was done in Ethiopia, the largest nation in the Horn of Africa. Ethiopia is the second most populous country in Africa and 11th in the world populous country. According to the United Nations Children’s Fund, there are 13 million under five years of age children in Ethiopia (26). The study was done from January-February 2024. This study used secondary data from the Ethiopia Mini Demographic Survey of 2019 (EMDHS). The EMDHS was a cross-sectional study conducted on identified areas defined as enumeration areas constituting geographic areas. It is national representative data that was collected through a community-based cross-sectional survey from March 21, 2019, to June 28, 2019. The 2019 EMDHS is the second EMDHS and the fifth DHS implemented in Ethiopia. The Ethiopian Public Health Institute (EPHI) conducted the survey in collaboration with the Central Statistical Agency (CSA) and the Federal Ministry of Health (FMoH), with technical assistance from ICF and financial as well as technical support from development partners(27).

### Population

This study focused on the children aged 12-35 months old that are sampled from all child vaccination-related data in Ethiopia. All 12-35 months of age children who have at least one confirmed vaccination history and whose vaccination card was seen at home or from health facilities were included in the study. The children were excluded if the dates of vaccination were inconsistent with their birthdates.

### Sample Size Determination and Sampling Techniques

The study included 1077 children aged 12-35 months who are vaccinated according to EPI and their collected data available in EMDHS 2019. The 2019 EMDHS utilized the census frame from the 2019 Ethiopia Population and Housing Census (EPHC), established by the Central Statistical Agency (CSA), consisting of 149,093 census enumeration areas (EAs). Each EA, on average, covers 131 households and contains data regarding EA location, urban or rural residency, and estimated residential household count. Ethiopia’s administrative structure was comprised of nine geographical regions and two administrative cities during the survey (27).

The sample design aimed at generating key indicators for the entire country, urban and rural areas individually, and for each of the nine regions and the two administrative cities. The sample selection process occurred in two stages within 21 strata, stratifying each region into urban and rural areas. In the first stage, 305 EAs (93 urban and 212 rural) were chosen with a probability proportional to EA size, while the second stage involved selecting a fixed number of 30 households per cluster using equal probability systematic selection from a newly established household listing. The survey carried out between March 21, 2019, and June 28, 2019, interviewed 8,855 women aged 15-49 from a nationally representative sample of 8,663 households, aiming to furnish estimates at national, regional, urban, and rural levels (27).

### Operational Definitions

#### Age-appropriate Vaccination

Age-appropriate vaccination is children’s vaccination timely within the WHO-recommended window period for basic vaccines: BCG (birth – 8 weeks), Penta1and OPV1 (6 weeks – 14 weeks); Penta2, and OPV2 (10 weeks – 18 weeks); Penta3, and OPV3 (14 weeks – 24 weeks)] and measles vaccine (9 months – 11 months) (28). Vaccination of children was coded as ‘1’ if the child received the vaccine in the recommended period, and coded as ‘0’ if vaccination was done before or after the recommended period. Age-appropriate vaccination was calculated from the summation of all “yes” scores for each specific basic vaccine type considered for this study.

According to the EMDHS report, the child must receive at least: one dose of BCG vaccine, which protects against tuberculosis; three doses of DPT-HepB-Hib, which protects against diphtheria, pertussis (whooping cough), tetanus, hepatitis B, and Haemophilus influenzae type b; three doses of polio vaccine; and one dose of measles vaccine (27). Therefore, we considered these vaccines in the analysis.

### Data Extraction Tools and Quality Control Methods

The data extraction checklist was adapted from the EMDHS to extract necessary data. The checklist was prepared in a way to extract socio-demographics, maternal and obstetric factors, and information-related factors from the large data. Before conducting any analysis, data was extracted depending on the inclusion criteria. Then variables were identified, recoded, cleaned, computed, and missing values were coded.

### Statistical Analysis

The extracted data was analyzed using the STATA version 14 statistical software. To account for differences in stratum selection and nonresponse probabilities, the data was weighted. Descriptive and inferential statistical methods were employed to present the data. Descriptive statistics such as frequency distributions and percentages were used.

A multi-level-logistic regression modeling was done to assess the association between the independent variables with the dependent variable. Variables with p-value <0.05 at 95% with AOR were declared as statistically significant using the me logit function. To check for multicollinearity among the independent variables, the variance inflation test was performed (a default function for STATA 14.0 version) and the final model was used by comparing model 0, model I, model II, and model III. In Model 0 the dependent variable was analyzed with the regional variable which means that the initial analysis was focused on how the dependent variable is influenced by the regional variable alone. In Model I the effect of an individual-level variable on a dependent variable was analyzed which means theanalysis lookedat how a specific variable at an individual level impacts the dependent variable. Model II examined the dependent variable and the community variable which is resident. Model III level examined using significant variables in both model I and model II by the stepwise analysis method. The model with the least AIC and BIC was used to identify model fitness.

### Ethical Considerations

For the data we requested www.DHSmeasure.org, to obtain permission to access the data set from https://www.dhsprogram.com/data/available-datasets.cfm by submitting abstract. The DHS public-use datasets are protected from identification of respondents, households, or sample communities by procedures approved by the Institution Review Board. The data files do not contain household addresses or names of individuals. The data set was not used for other purposes except for the intended objective.

## Result

### Socio-Demographic Characteristics

A total of 1077 children aged 12-35months whose vaccination card was checked at home or health facility in MEDHS 2019 were included.The mean age of mothers was 28.58 ±6.27 with a minimum of 17 years and a maximum of 49 years old. Similarly, the mean age of children was 22.34±6.70 with a minimum of 12 months and a maximum of 35 months. About half of the children were male 549 (50.97) (Table 1).

**Table 1:** Distribution of socio-demographic characteristics (n=1077).

| **Variables** | **Categories** | **Frequency** | **Percent** |
| --- | --- | --- | --- |
| Region | Tigray | 134 | 12.44 |
|  | Afar | 52 | 4.83 |
|  | Amhara | 117 | 10.86 |
|  | Oromia | 135 | 12.53 |
|  | Somali | 51 | 4.74 |
|  | Benishangul Gumuz | 114 | 10.58 |
|  | SNNPR | 94 | 8.73 |
|  | Gambela | 77 | 7.15 |
|  | Harari | 85 | 7.89 |
|  | Addis Ababa | 110 | 10.21 |
|  | Diredawa | 108 | 10.03 |
| Place of residence | Urban | 357 | 33.15 |
|  | Rural | 720 | 66.85 |
| Age of the mothers | 15-19 | 39 | 3.65 |
|  | 20-24 | 218 | 20.24 |
|  | 25-29 | 390 | 36.21 |
|  | 30-34 | 218 | 20.24 |
|  | 35-39 | 135 | 12.53 |
|  | >=40 | 77 | 7.15 |
| Mother educational status | No education | 464 | 43.08 |
|  | Primary | 399 | 37.05 |
|  | Secondary | 120 | 11.14 |
|  | Higher | 94 | 8.73 |

| Variable | Category | Frequency | Percent |
| --- | --- | --- | --- |
| Religion | Orthodox | 418 | 38.81 |
|  | Protestant | 181 | 16.81 |
|  | Muslim | 463 | 42.99 |
|  | Other * | 15 | 1.39 |
| Sex of HH<sup>++</sup> head | Male | 888 | 82.45 |
|  | Female | 189 | 17.55 |
| Wealthy status | Poor | 389 | 36.12 |
|  | Middle | 167 | 15.51 |
|  | Rich | 521 | 48.38 |
| Marital status | currently in union/living with a man | 1025 | 95.17 |
|  | Other ** | 22 | 4.83 |
| Family size | Less than five | 376 | 34.91 |
|  | Above 5 | 701 | 65.09 |
| Number of <5 children at home | <=1 | 532 | 49.40 |
|  | >=2 | 545 | 50.60 |
| Sex of children | Male | 549 | 50.97 |
|  | Female | 528 | 49.03 |
| Age of children | Between 12-24 months | 645 | 59.89 |
|  | Above 24 months | 432 | 40.11 |

\*=Catholic, Traditional, \*\*= Single, widowed, Divorced

### Maternal and obstetric-related Factors

The study revealed that almost all, 1056(98.05) births were singleton. Concerning ANC follow-upa bit higher than half, 504(55.96%) of mothers had 4 or more ANC visits. The study revealed that 745(69.17%) of mothers delivered in health facilities, while about two-thirds 456 (66.47%) of mothers had PNC follow (Table 2).

**Table: 2.** Distribution of maternal and Obstetric characteristics (n=1077).

| Variable | Category | Frequency | Percent |
| --- | --- | --- | --- |
| Place of delivery | Health facility delivery | 745 | 69.17 |
|  | Home delivery | 332 | 30.83 |
| Birth order of the child | First | 270 | 25.07 |
|  | 2-4 | 492 | 45.68 |
|  | >=5 | 315 | 29.25 |
| Birth in the past 5 years | One | 637 | 59.15 |
|  | Two | 384 | 35.65 |
|  | Three or four | 56 | 5.20 |
| Type of birth | Single birth | 1056 | 98.05 |
|  | Multiple births | 21 | 1.95 |
| ANC visit | No ANC visits | 117 | 12.12 |
|  | Less than 4 ANC visits | 308 | 31.92 |
|  | >=4 ANC visits | 504 | 55.96 |
| PNC | No | 230 | 33.53 |
|  | Yes | 456 | 66.47 |

NB: ANC= Antenatal Care, PNC= Post Natal Care

### Media Exposure

The study revealed that more than half, 582 (54.04%) of the children’s mothers had no access to media exposure (Fig.1).

**Figure 1.**
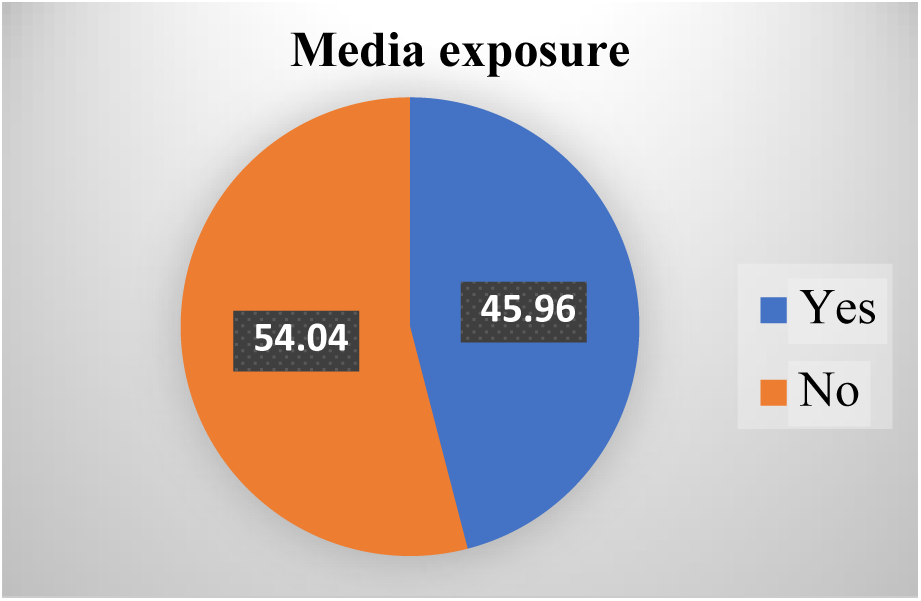
Distribution of children’s mothers’ exposure to media

### Prevalence of age-appropriate Vaccination

The prevalence of age-appropriate vaccination was 21.17% 95%CI (18.73-23.61) of children who received the basic vaccinations at an appropriate age. Furthermore, theresults of the study showed that the majority of children were vaccinated for all basic vaccinations as presented in Table 3.

**Table 3:** Distribution of age-appropriate vaccination among children aged 12-35 months.

| Basic Vaccinations | Response | Appropriate time |  |
| --- | --- | --- | --- |
|  |  | Frequency | Percent |
| BCG | Yes | 607 | 62.13 |
|  | No | 370 | 37.87 |
| OPV1 | Yes | 734 | 69.71 |
|  | No | 319 | 30.29 |
| Opv2 | Yes | 678 | 68.55 |
|  | No | 311 | 31.45 |
| OPV3 | Yes | 662 | 71.49 |
|  | No | 264 | 28.51 |
| Penta1 | Yes | 733 | 69.22 |
|  | No | 326 | 30.78 |
| Penta 2 | Yes | 697 | 68.98 |
|  | No | 309 | 31.02 |
| Penta 3 | Yes | 670 | 72.28 |
|  | No | 257 | 27.72 |
| Measles | Yes | 473 | 60.80 |
|  | No | 305 | 39.20 |

### Factors Associated With Age-appropriate Vaccination Among Children Aged 12-35months

In the multilevel logistic regression analysis, the age of mothers, educational status of the mothers, and childbirth in the past five years were statistically significant determinants of age-appropriate vaccination

The study identified that children’s mothers aged >= 40 years were 4.05 times (AOR=4.05 at 95%CI1.03, 15.83) more likely to vaccinate their child at an appropriate age compared to those aged between 15-19 years old. Similarly, 35-39-year-old mothers were 4.62 times (AOR= 4.62 at 95%CI 1.27, 16.71) more likely to vaccinate their child at an appropriate age compared to those aged between 15-19 years old. In addition, 25-29-year-old mothers were 4.07 times (AOR =4.07 at 95%CI1.18, 14.03) more likely to vaccinate their child at appropriate age compared to those aged between 15-19 years old.

Regarding maternal education, mothers who completed secondary education were 1.85 times (AOR=1.85 at 95% CI1.06, 3.22) more likely to vaccinate their child at an appropriate age compared to those who had no education. In addition, mothers who completed primary education were 1.60 times (AOR= 1.60, 95% CI=1.07, 2.41) more likely vaccinated at an appropriate age compared to those who had no education.

Regarding place of residence, mothers wholive in rural areas were 66% (AOR=.34 at 95%CI.23,.51) less likely to vaccinate their child at an appropriate age compared to those who live in urban areas (Table 4).

**Table 4:**
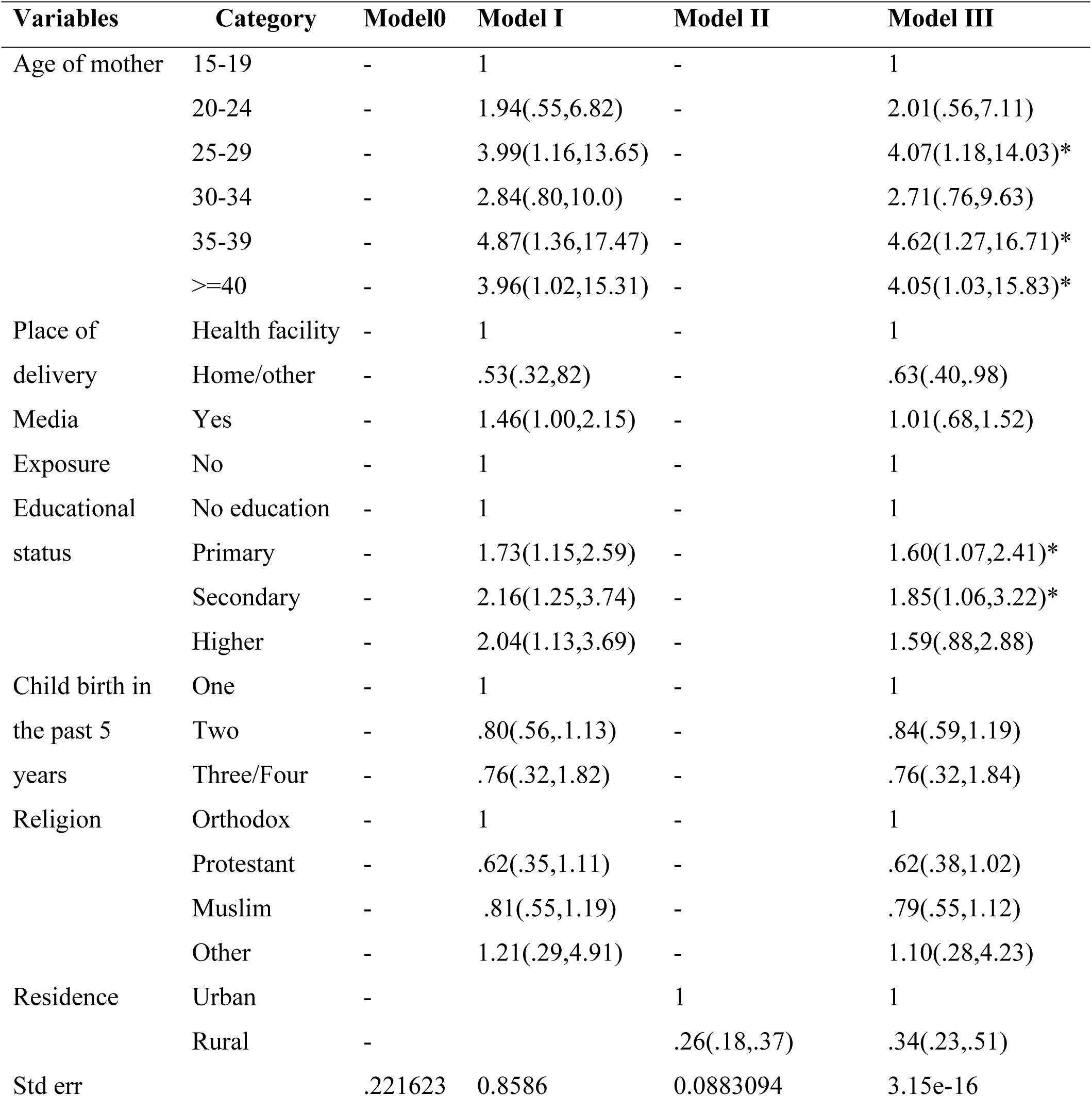
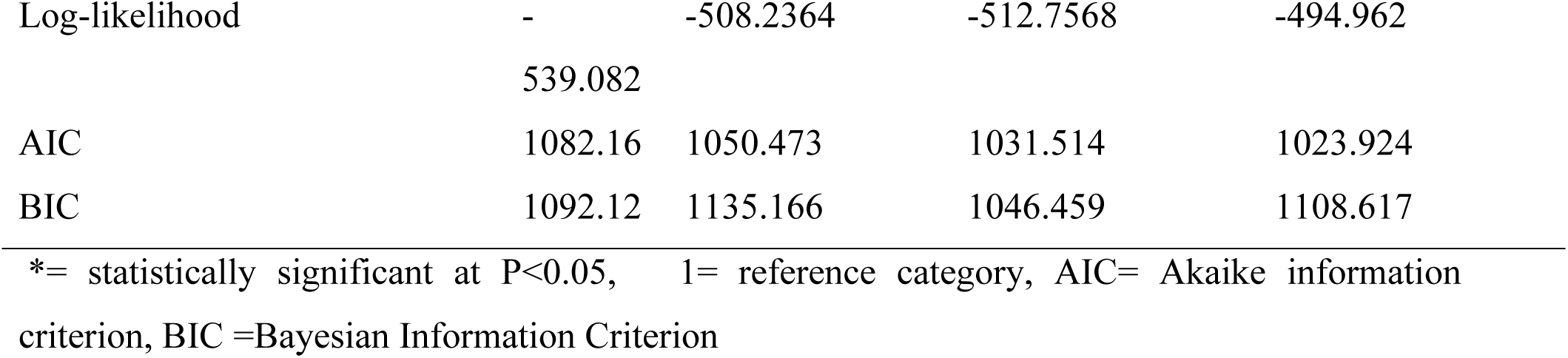
Multilevel logistic regression analysis of factors associated with age-appropriate vaccination among children aged 12-35 months.

### Model fit statistics

As shown in Table 4 below, a small number of AIC and BIC was Model IV, which indicates that the model well explained the factors better than the Null Model, Models II and III. Therefore, this made the final as the best-fitted model than others.

## Discussion

The present study aimed to assess the magnitude and factors associated with age-appropriate vaccination among children aged 12-35 months in Ethiopia. Our results showed thatthe prevalence of age-appropriate vaccination in this study was 21.17%. Mother’s age educational status, and residence rural were significant predictors of age-appropriate vaccinations.

According to the findings from the current study age-appropriate vaccination coverage is very low (21.17 %) compared to the crude prevalence of child vaccination in Ethiopia. This finding is not supported by comparable literature where this figure is very low. The majority of the studies on this area are small samples and addressed children from limited study areas. Consequently, the age-appropriate vaccination coverage in this study is lower than the prevalence of study findings in El Salvador(26.7%) (29). Similarly, this finding is lesser than the timely vaccination of specific vaccine types in India where the coverage is 31%, and 34% for BCG and MCV (30). Generally, the very low finding of age-appropriate vaccination could be related to low economic status, low education, and other factors in Ethiopia (31–33).

In the present study, children of mothers aged 25-29, 35-39, and 40 and above were more likely to be vaccinated at an appropriate age compared to the children of mothers 15-19 years old during the data collection. This indicates that older mothers are at a better oddsof vaccinating their children in Ethiopia. This result is supported by the study conducted in China (34), and in Tanzania where younger mothers are at a risk of low age-appropriate vaccination (35). This could be because older mothers can have valuable experience in child caring they have likely raised children before, and they can have more exposure to health facilities and health information that can be delivered there (36, 37). On the other hand, mothers aged above 35 years were less likely to vaccinate their children on time (38). Scientific research established that both extremities mother’s age affects child caring (37).

Moreover, children of more educated mothers were more likely to be vaccinated at appropriate age. The children of mothers who have primary, secondary, and higher education had two times higher odds of being vaccinated compared to children from mothers who did not attend formal education. This study finding is supported by the study from China (34), Saudi Arabia (39), Tanzania (35), Cameron (40), and Ethiopia (41). This significance among educated mothers may be because of educated mothers being able to have awareness of the benefits of child vaccination. Also, educated mothers can be better at understanding the health care providers’ counseling and messages on vaccination during awareness creation sessions at health facilities and at the community level many times the mothers get childcare information from health extension workers and women development armies in Ethiopia (42, 43). In support of this, scientific research has confirmed that children of educated mothers have a higher rate of complete immunization. This is because maternal education leads to better literacy skills and health-seeking behavior, which in turn improves immunization uptake for their children (44, 45).

Furthermore, children of mothers who live in rural areas were less likely vaccinated at the appropriate age compared to those who live in rural areas. This finding is supported by other studies in Nigeria(46) and Kenya(47). This could be due to various factors like limited access to health care services, inadequate infrastructure, and lower educational levels in rural settings. This implies that the government has to look into another strategic direction like community engagement and targeted intervention mechanisms to minimize rural-urban disparities.

Despite the current analysis being based on the most recent nationally representative survey data and usinga multilevel logistic regression to handle the variation within the level due to the hierarchical nature of the EMDHS data, it is not free of limitation. One of the limitations of the study is the difficulty of showing temporal association because of the cross-sectional nature of the study. Another limitation, since the EMDHS survey is based on the participants’ reports, there might be a recall bias.

## Conclusion

From this study, we concluded that the prevalence of age-appropriate vaccination of children in Ethiopia is significantly below the desired level. Factors such as mother’s age, education, and residence were significant predictors of age-appropriate vaccinations. Hence, we recommend that the stakeholders should give due attention to promote timely vaccination of children for their age.

## Data Availability

The data used for this study is included within the article itself

## Acknowledgment

The authors would like to express gratitude to www.DHSmeasure.orgfor granting access to the datasets.

